# Assessment of Tear film Stability and Ocular Surface Disease Index among Glaucoma Subjects living with and without Diabetes Mellitus in Owerri, Imo State, Nigeria

**DOI:** 10.64898/2026.08.31.26361784

**Authors:** Chigozie Mary Chukwuoha, Emmanuel C. Esenwah, Nwakaego C. Ikoro, Godwin-Ovenseri-Ogbomo, Young Christian Azuamah, Nkiru Euphresia Odimegwu, Anthony U. Megwas, Genevieve Ugwoke, Jacqueline E. Obioma-Elemba, Yadirichukwu Eronini, Ezinne Chinenye Nkeremuzor

**Author notes:** **Corresponding Author:** Chigozie Mary Chukwuoha, **E-mail:**. **Author E-mails** Godwin-Ovenseri-Ogbomo.

## Abstract

**Objective:** The ocular surface is an integral unit of the visual system. Its smooth and wet surface which is maintained by the tear film, is the most important refractive interface of the visual system. This study compared ocular surface alterations among glaucoma subjects living with and without diabetes mellitus in a Owerri, Imo State, Nigeria

**Methods:** Subjects diagnosed of glaucoma living with and without diabetes mellitus were included in the study. Ocular surface assessment included the Ocular Surface Disease Index (OSDI) questionnaire, and tear break-up time (TBUT). Descriptive statistics, independent samples t-test and Chi-square test were used to examine the data at 0.05 level of significance.

**Results:** Considerable shorter tear break-up time (11.3 ± 9.0s) was seen among glaucoma subjects living with diabetes mellitus than those living without diabetes mellitus (16.0 ± 10.6s; p = 0.003), demonstrating the higher tear film instability. This was similarly reflected by the mean OSDI score, which was considerably higher in glaucoma patients living with diabetes mellitus (40.0 ± 23.4) compared with those without diabetes mellitus (28.1 ± 21.8; p = 0.001) showing a larger ocular surface symptom load. However, although the frequency of severe OSDI symptoms was greater in glaucoma subjects living with diabetes mellitus, the relationship between diabetes status and OSDI severity categories was not statistically significant (χ 2 = 7.584, p = 0.055).

**Conclusion:** Diabetes mellitus significantly impacts the health of the ocular surface in glaucoma subjects. The combination of glaucoma and diabetes may result in more rapid tear film instability, increased ocular pain, and decreased quality of life and adherence to long-term glaucoma medication.

## 1. Introduction

The ocular surface is an integral unit of the visual system. Its smooth and wet surface which is maintained by the tear film, is the most important refractive interface of the visual system.^1, 2^ The cornea and ocular surface function as major barriers and the eye’s primary refractive medium, which require accurate coordination to maintain transparency, structural integrity, and immune protection.^3^ This smooth and wet surface enhances the transmission of light through the ocular media to the retina for photoreceptor stimulation.^1,4^ Being always exposed to environmental stressors, this interface relies on the stability of the tear film, epithelial architecture, mucin layers, and limbal stem cells to maintain its role. Any alterations in these component can lead to vision impairment and make the ocular sruface prone to pathologic conditions.^2,3,5^

The array of proteins, glycoproteins and lipids in tears function to maintain a stable, well-lubricated and smooth optical surface.^3,6^ The tear film is a thin fluid layer covering the ocular surface. It is the interface of the ocular surface with the environment and is responsible for ocular surface comfort, mechanical, environmental and immune protection and epithelial (both corneal and conjunctival) health. It forms smooth, refracting surface for vision.^7-9^

Homeostatic balance leads to stability of the tear film, which makes it possible to carry out its functions as lubrication, nutrition and protection of ocular surface.^9,10^ Nonetheless, this stability can be disturbed in tear film layers’ deficiencies, defective spreading of the tear film, in some general diseases and during application of some systemic and/or topical medications and dry eye disease evolve as a consequence.

Glaucoma is a complex eye condition characterized by elevated intraocular pressure (IOP) that may progress to vision loss over time. It is the second leading cause of permanent blindness worlwide.^11,12^ It can be categorized into either primary or secondary types and further into open-angle or closed-angle variants within each type of glaucoma. Adult glaucoma includes primary open-angle glaucoma (POAG) and angle-closure glaucoma, as well as secondary open and angle-closure glaucoma,^13^ with a specific focus on the most prevalent type, POAG.^12,14^

Reduction of intraocular pressure is the only proven method to effectively manage glaucoma as a disease. This can be usually achieved by anti-glaucoma medications, laser therapy, or surgical intervention.^15^ While lowering the intraocular pressure (IOP) is the most important measure to prevent further damage to the optic disc, long-term topical treatment represents a continuous hazard to the ocular surface homeostasis. Long term topical glaucoma therapy has been associated with reduced density of goblet cells and squamous metaplasia of the conjunctival epithelium,^16,17^ dysfunction of meibomian glands, conjunctival and corneal desquamation ^16^ and overexpression of proinflammatory cytokines.^17,18^ Significant loss of goblet cells, which can cause dry eye, inflammation, and fibrosis, was observed in animal and human models.^16,17^ As a consequence of the inflammatory changes, chronic use of IOP-lowering medications can also affect bleb scarring in filtration surgery, since it is a risk factor for conjunctival fibrosis which can ultimately result in failure of trabeculectomy.^17,19^

Some studies have investigated the coexistence of glaucoma and ocular surface disease (OSD).^17,20,21^ The prevalence of OSD is estimated to vary between 5% and 30% in the general population, but may increase up to 50% in glaucoma patients under medical treatment.^2,17^ Several risk factors such as aging, hormone imbalance, systemic comorbidities, systemic medications and environmental exposure are frequently associated with the chronic use of IOP-lowering eye drops, triggering proinflammatory responses and ocular surface dysfunction in patients with glaucoma.^22^

Diabetes is one of the common causes of blindness in persons aged 20-70 years. Cataract and retinopathy are well known ocular complications of diabetes. However, in recent times, attention has been drawn to ocular surface problems, especially dry eye in diabetic patients.^23^ Studies have revealed that diabetics are more prone than the overall population to suffer from dry eyes, owing to various reasons. According to the International Diabetes Federation, over half of diabetics have dry eye. Also, data from a population based study showed that individuals with diabetes had thicker cornea.^24,25^

Abnormalities in tear secretion, alteration of epithelial barrier and autonomic neuropathy lead to tear film and ocular surface changes in diabetes, thus causing dry eye. Much of our current understanding of ocular complications of DM have focused primarily on the retina, yet, the ocular surface is an important structure impacted by diabetes.^26^ An intimate relationship between dry eye and diabetes mellitus (DM) with changes on the ocular surface is therefore suggested. This study aimed to comparatively assess tear film stability and ocular surface disease symptom among glaucoma subjects living with and without Diabetes Mellitus in a South-Eastern Nigeria Population by assessing the Tear Break Up Time (TBUT) and Ocular surface Disease Index (OSDI) score.

## 2. Material and Methods

### 2.1 Research design

Data used in study were drawn from a larger clinic-based cross-sectional study investing the comparative assessment of ocular surface changes among glaucoma subjects living with and without diabetes mellitus in owerri, Imo State, Nigeria.

### 2.2 Population of study

The study population comprised 157 patients who gave informed consent and met with the inclusion criteria for this study, diagnosed with glaucoma and living with or without diabetes, who visited an eye care facility in Owerri, Imo State, Nigeria.

### 2.3 Inclusion criteria

- Subjects diagnosed of open angle-glaucoma who gave consent to participate in the study
- Glaucoma patients living with or without diabetes as comorbidity.
- Those on topical antiglaucoma medication
- No age limitation

### 2.4 Exclusion criteria

Patients with history of ocular trauma, infection or deformities in the external eye, those who have undergone ocular surgery or laser procedure 6 months prior to the study, presence of corneal pathologies, contact lens use, chronic use of other topical medication, systemic diseases such as rheumatoid arthritis, Sjogren’s syndrome, thyroid disease, and lupus, exposure to dry climate smoke and continuous computer screen users.

### 2.5 Procedure for data collection

This included detailed case history to obtain required information such as data such as age, gender, duration of glaucoma and diabetes, use of topical antiglaucoma medication, previous ocular surgeries and other ocular and systemic comorbidities.

#### 2.5.1 Ocular Surface Disease Index

The ocular surface was examined and OSD symptoms were evaluated using the Ocular Surface Disease Index (OSDI) questionnaire (developed by Allergan), which is the most frequently used survey instrument for assessment of ocular surface disease severity.^17,27^ The OSDI was assessed on a scale of 0 to 100, with higher scores showing greater instability.

The OSD symptoms were scored from 0 (indicating symptoms none of the time) to 4 (symptoms all the time) and the total score ranged from 0 to 100 and calculated using the formula below:

*Sum of scores for all questions answered (SSQA) x 100/no. of questions answered (nQA) x 4*.

The scores were graded as follows:

0 to 12 = normal,

13 to 22 = mild dry eye disease,

23 to 32 = moderate dry eye disease; 33 or above = severe dry eye disease.

Ocular surface parameters were evaluated using the following procedures:

#### 2.5.2 Tear Break-up Time

Fluorescein tear breakup time (FTBUT) test was used to determine the stability of the tear film. TBUT is the time interval between the last blink and tear film disruption. This method of staining the ocular surface with fluorescein to assess tear breakup time is a subjective assessment of tear film stability and is measured in seconds.

A standard fluorescein strip was applied to the lower fornix without topical anesthetic. The patient was asked to blink several times to spread the dye well, and the tear film layer was observed under wide illumination using a cobalt blue filter on a slit lamp biomiscroscope. At the first appearance of the dry spot(s), the stopwatch was click-stopped and the time interval between the last blink and the first appearance of the dry spot(s) was recorded in seconds as the TBUT. Values gotten were graded as follows:

<5s - severe instability/evaporative dry eye disease

5-10s - mild-to-moderate instability/dry eye disease

>10s - normal

### 2.6 Statistical analysis

Data were analyzed using the IBM Statistical Package for the Social Sciences (SPSS) version 31. Quantitative variables were described using mean and standard deviation (SD) for continuous variables whereas categorical variables were presented using frequencies and percentages. Independent sample t-test was used to determine the difference between the two groups. The chi-square test was used to analyze categorical variables.

Correlation analysis was used to assess the relationship between the variables in the study. A significance level (*p value*) of ≤ 0.05 was considered statistically significant for all variables in the study.

### 2.7 Ethical Approval/Informed Consent

Ethical Approval was obtained from the Ethics Committee, School of Health Technology, Federal University of Technology, Owerri, to carry out this study after review, on the 8^th^ of July, 2025. No specific protocol number was assigned by the committee. Informed consent was also gotten from subjects before inclusion in the study.

## 3. Results

The study population comprised 48 (30.6%) males and 109 (69.4%) females with a mean age of 62.9±11.1. Figure I below shows the distribution of glaucoma subjects living with and without diabetes mellitus in Owerri according to age. Among the subjects included in the study population, 74 (47.1%) with a mean age of 66.9±7.3, made up the population of glaucoma subjects living with DM, while 83 (52.9%) with a mean age of 59.2±12.5, made up the population of glaucoma subjects living without DM.

**Figure 1:**
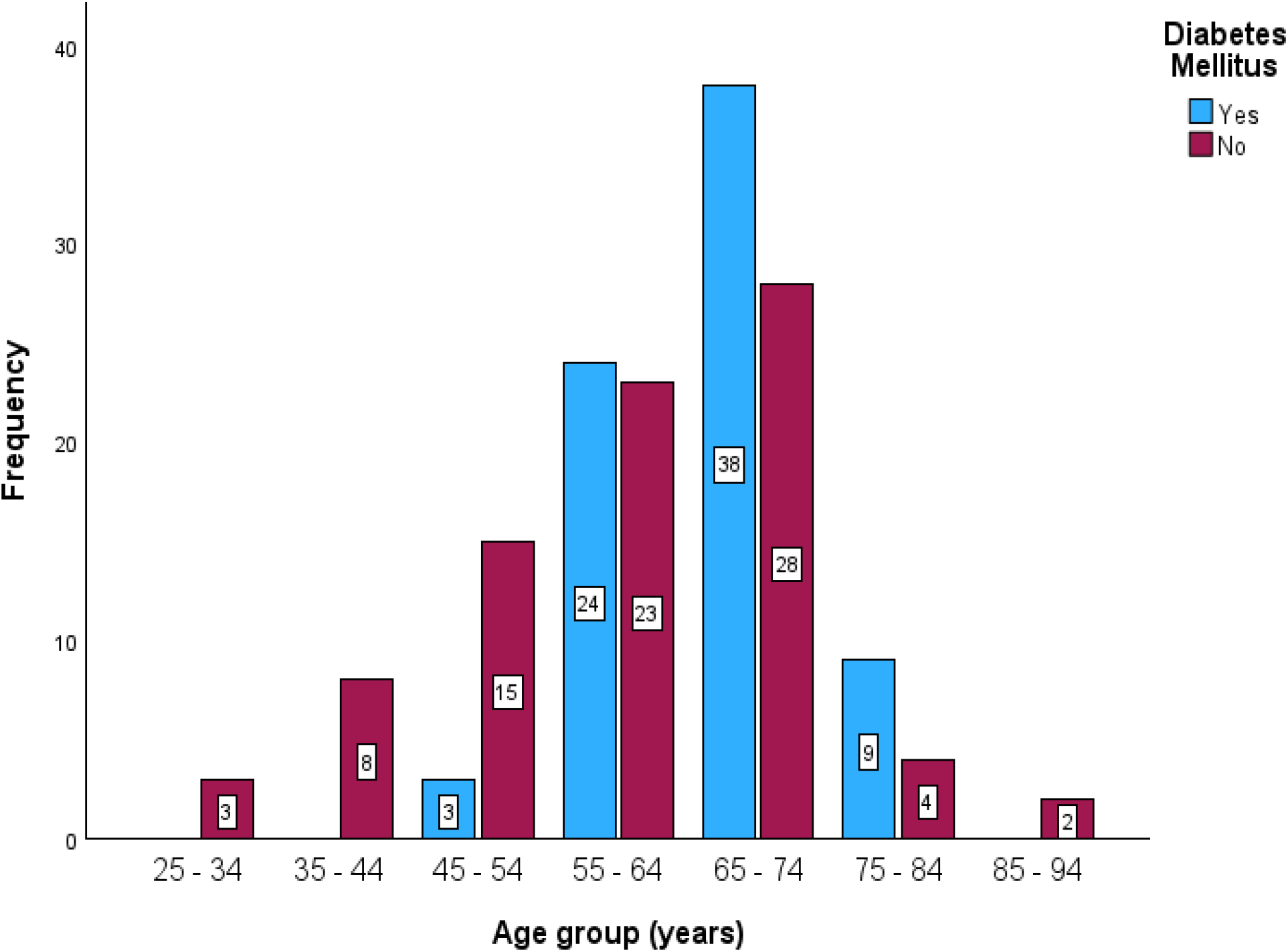
Distribution of Glaucoma subjects living with and without DM in Owerri according to Age

Based on the age groups, the highest population of glaucoma subject living with DM was seen in the age group 65-74 years (38, 51.4%). This was followed by the 55-64 years age group (24, 32.4%) and 75-84 years (9, 12.2%). The least population of glaucoma subjects living with DM was seen in the 45-54 years age group (3, 4.1%). No glaucoma subjects living with DM were seen among the age groups 25-34 years, 35-44years and 85-94 years.

Among the glaucoma subjects living without DM, the highest population of subjects was seen among the 65-74 years age group (28, 33.3%). This was followed by the age group 55-64 years (23, 27.1%) 45-54 years (15, 18.1%), 35-44 years (8, 9.6%), 75-84 years (4, 4.8%) and 25-34 years (3, 3.6%). The least population was seen in the 85-94 years age group (2, 2.4%).

### Distribution of glaucoma subjects living with and without DM in Owerri according to Sex

Based on sex, 21 (28.4%) of the male subjects with a mean age of 65.3±6.5, and 53 (71.6%) of the female subjects with a mean age 67.6± 7.6, comprised the population of glaucoma subjects living with DM. For glaucoma subjects living without DM, 27 (56.3%) of the male subjects with a mean age of 63.1±13.6, and 56 (51.4%) of the female subjects with a mean age of 57.3±11.7, made up the population of study.

**Table 1:** Distribution of Glaucoma subjects living with and without DM in Owerri according to Sex.

| Sex | Glaucoma subjects living with DM | % | Glaucoma subjects living without DM | % | Total % |
| --- | --- | --- | --- | --- | --- |
| Male | 21 | 28.4 | 27 | 32.5 | 30.6 |
| Female | 53 | 71.6 | 56 | 67.5 | 69.4 |
| **Total** | **74** | **100** | **83** | **100** | **100** |

### Tear Break Up Time (TBUT) amomg glaucoma subjects living with and without DM in Owerri

The mean TBUT among all subjects incuded in this study was 13.8± 10.1. For glaucoma subjects living with DM in Owerri, the mean TBUT was 11.3± 9.0, while for glaucoma subjects living without DM, the mean TBUT was 16.0±10.6. Overall, 68.8% of the subjects in this study had TBUT above 8 seconds. This was followed by 22.9% of the population with TBUT between 5-7 seconds. The least TBUT was seen among 8.3% of the study population, who had TBUT less than 5 seconds.

Among the population of glaucoma subjects living with DM in Owerri, 56.8% had had TBUT above 8 seconds. This was followed by 37.8% of subjects in this group, with TBUT between 5-7 seconds, while the least TBUT was seen among 5.4% of glaucoma subjects living with DM, who had TBUT less than 5 seconds.

For glaucoma subjects living without diabetes mellitus in Owerri, 79.5% had TBUT above 8 seconds, 9.6% had TBUT between 5-7seconds, while the least time was seen among 10.8% of subjects in this group, who had TBUT less than 5seconds.

**Table 2:** TBUT among glaucoma subjects living with and without diabetes mellitus in Owerri.

| TBUT (secs) | Glaucoma subjects living with DM | % | Glaucoma subjects living without DM | % | Total | % Total |
| --- | --- | --- | --- | --- | --- | --- |
| ≥8 (Normal) | 42 | 56.8 | 66 | 79.5 | 108 | 68.8 |
| 5-7(mild-moderate) | 28 | 37.8 | 8 | 9.6 | 36 | 22.9 |
| <5 (severe) | 4 | 5.4 | 9 | 10.8 | 13 | 8.3 |
| **Total** | **74** | **100** | **83** | **100** | **157** | **100** |

### Ocular Surface Disease Index (OSDI) among glaucoma subjects living with and without Diabetes Mellitus in Owerri

The mean OSDI score among all subjects included in this study was 33.7± 23.3. For glaucoma subjects living with DM in Owerri, the mean OSDI score was 40.0± 23.4, while for glaucoma subjects living without DM, the mean OSDI score was 28.1±21.8. Overall, the highest OSDI score was seen among 45.2% of the population of study, with score ranging from 33-100. This was followed by 22.3%, with OSDI score ranging from 0-12 and 19.7% with OSDI score ranging from 23-32. The least OSDI score was seen among 12.7% of the population, with OSDI score ranging from 13-22.

Among glaucoma subjects living with DM in Owerri, the highest OSDI score was seen among those with score between 33-100 (56.8%), followed by those with OSDI score between 0-12 (17.6%) and those with OSDI score between 23-32 (16.2%). The least OSDI score was seen among those with OSDI score between 13-22 (9.5%).

For glaucoma subjects living without DM in Owerri, the highest OSDI score was seen among those with score between 33-100 (34.9%), followed by those with OSDI score between 0-12 (26.5%) and those with OSDI score between 23-32 (22.9%). The least OSDI score was seen among those with OSDI score between 13-22 (15.7%).

**Table 3:** OSDI among Glaucoma subjects living with and without Diabetes Mellitus in Owerri.

| OSDI score | Glaucoma subjects living with DM | % | Glaucoma subjects living without DM | % | Total | % Total |
| --- | --- | --- | --- | --- | --- | --- |
| 0 – 12 (normal) | 13 | 17.6 | 22 | 26.5 | 35 | 22.3 |
| 13 – 22 (mild) | 7 | 9.5 | 13 | 15.7 | 20 | 12.7 |
| 23 – 32 (moderate) | 12 | 16.2 | 19 | 22.9 | 31 | 19.7 |
| 33 – 100 (Severe) | 42 | 56.8 | 29 | 34.9 | 71 | 45.2 |
| **Total** | **74** | **100** | **83** | **100** | **157** | **100** |

### Difference in Tear Break Up Time among Glaucoma subjects living with and without Diabetes Mellitus in Owerri

A statistically significant difference (*p-value* = .003) at 0.05 significance level, was found in the mean TBUT between glaucoma subjects living with and without DM in Owerri. In terms of grading, the Chi square analysis showed a statistically significant relationship between diabetes status and grading of TBUT, χ^2^ = 17.91, *p*<.001. Hence, diabetes status was significantly associated with the measured time and grading of TBUT among the glaucoma subjects included in the study.

**Table 4:** Summary of data analysis for difference in the Tear Break Up Time among Glaucoma subjects living with and without Diabetes Mellitus in Owerri.

| Tear Break Up Time |  |  |
| --- | --- | --- |
| Glaucoma subjects living with DM | Glaucoma subjects living without DM | $P$-value |
| 74 | 83 | .003 |
| Chi square $\chi^2 = 17.91$, $p < .001$ | | |

### Difference in Ocular Surface Disease Index among Glaucoma subjects living with and without Diabetes Mellitus in Owerri

A statistically significant difference (*p* = 0.001) at 0.05 significance level was seen in the mean OSDI between glaucoma subjects living with and without DM in Owerri. In terms of grading, the chi square analysis showed no statistically significant relationship between diabetes status and grading of OSDI, χ^2^ = 7.584, *p*=.055.

Hence, diabetes status was not significantly associated with the grading of OSDI among the glaucoma subjects included in the study.

**Table 5:**
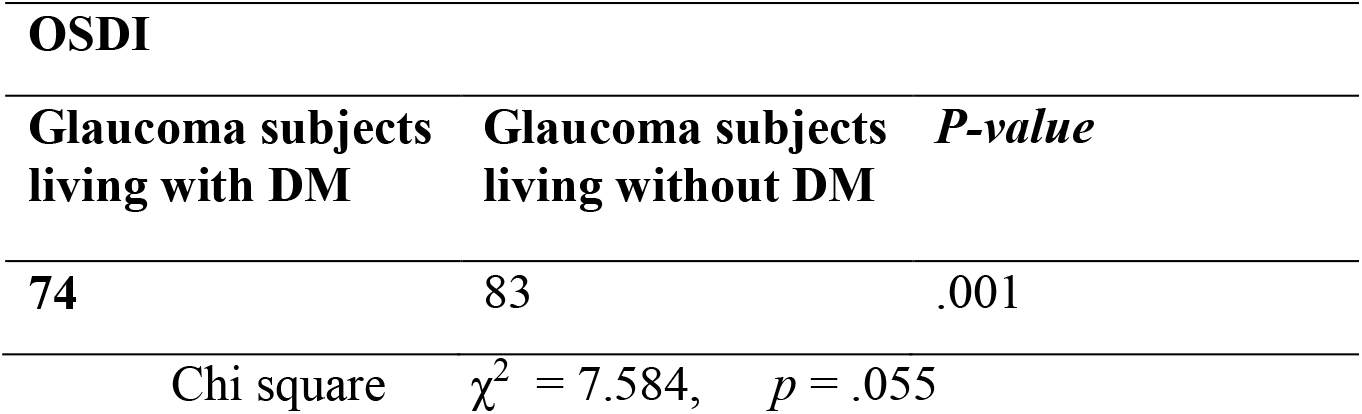
Summary of data analysis for difference in the Ocular Surface Disease Index among Glaucoma subjects living with and without Diabetes Mellitus in Owerri.

| OSDI |  |  |
| --- | --- | --- |
| Glaucoma subjects living with DM | Glaucoma subjects living without DM | P-value |
| 74 | 83 | .001 |
| Chi square $\chi^2 = 7.584,$ $p = .055$ | | |

## 4. Discussion

The older age distribution of individuals with glaucoma and diabetes has substantial consequences for ocular surface health. Older age is independently related with diminished lacrimal gland function, meibomian gland dysfunction, decreased corneal sensitivity, loss of goblet cell and tear film instability. These age-related alterations together with diabetes mellitus and prolonged topical antiglaucoma medication may predispose patients to more severe ocular surface illness, which is particularly pertinent to the later objectives of the present study.

Sex distribution showed the entire study population comprised 69.4% females and 30.6% men. Similarly, women formed the highest group of glaucoma patients living with diabetes mellitus (71.6%) and without diabetes mellitus (67.5%). This gender preponderance may not necessarily indicate that women are more biologically susceptible to glaucoma but may rather be due to variations in healthcare utilisation and health-seeking behaviour. Generally, women are more likely than males to use preventive healthcare services, visit routine eye clinics, and engage in health research, resulting in a higher representation in hospital-based studies.^28,29^

In the present study, the female glaucoma patients with diabetes were likewise, somewhat older on the average than the male glaucoma patients with diabetes. This result may also be explained by the fact that women have a higher life expectancy, and are thus cumulatively exposed to more chronic systemic disorders such as diabetes mellitus and glaucoma. In addition, it has been shown that postmenopausal hormonal changes affect ocular blood flow, neuroprotection and ocular surface homeostasis, although current data is unclear on their direct role in the development of glaucoma.^30^

In general, the demographic results showed that glaucoma patients with co-existing diabetes mellitus were older than those without diabetes and females were the predominant participants in both research groups. These discoveries are of therapeutic relevance since advanced age and diabetes mellitus are independently related with ocular surface dysfunction, tear film homeostasis and dry eye illness. Thus, the demographic profile of the present study implies that glaucoma patients with diabetes, especially the older population, may have higher susceptibility to ocular surface alterations than glaucoma patients without diabetes. This gives an essential context for the interpretation of later data on tear break-up time and Ocular Surface Disease Index.

In the current study, mean tear break-up time (TBUT) was shorter in glaucoma subjects living with diabetes mellitus compared to glaucoma subjects living without diabetes mellitus indicating worse tear film stability in the diabetic glaucoma group. This is consistent with the systematic review and meta-analysis by,^31^ who found that diabetic patients had significantly shorter invasive TBUT than non-diabetic controls, suggesting significantly impaired tear film stability in people living with diabetes.

Another systematic review and meta-analysis reported that, in the majority of empirical studies, patients with type 2 diabetes mellitus had significantly shorter TBUT than non-diabetic controls, with few studies finding no statistically significant difference owing to methodological heterogeneity.^32^

This results of this study is also corroborated by glaucoma-specific investigations as seen in a Nigerian study which reported that the prevalence of aberrant TBUT was considerably higher in glaucoma patients on long-term topical antiglaucoma drugs compared with age and sex-matched controls (p < 0.001). This suggests that glaucoma and its treatment negatively impair tear film stability. ^33^ Similarly, some other study observed that TBUT was considerably lower in glaucoma patients on persistent topical antiglaucoma drugs compared to healthy controls (p = 0.003) indicating that glaucoma treatment is related with tear film instability. (Ray et al., 2023)

In terms of Ocular Surface Disease Index (OSDI), the mean score was higher in the glaucoma subjects living with diabetes mellitus (40.0 ± 23.4) than the glaucoma subjects living without diabetes mellitus (28.1 ± 21.8) suggesting more severe ocular surface symptoms among glaucoma subjects with diabetes. Furthermore, a higher percentage of glaucoma subjects living with diabetes (56.8%) reported severe OSDI scores (33–100) than those without diabetes (34.9%), suggesting that diabetes may exacerbate symptoms of ocular surface illness. This finding concurs with a previous Nigerian study which found a significantly higher OSDI score among glaucoma patients on chronic topical anti-glaucoma medications than healthy controls (P<0.001),^33^ indicating a significant burden of ocular surface symptoms due to chronic glaucoma therapy. Significantly higher OSDI scores in medically treated glaucoma patients than in healthy controls, while patients who had undergone trabeculectomy showed relatively better ocular surface status, suggesting that long-term use of topical medication worsens ocular surface symptoms have also been revealed by some authors.^34^

Similar results were reported in Ethiopia among glaucoma patients on topical hypotensive drugs who were assessed using OSDI questionnaire along with TBUT, Schirmer test and corneal staining. Dry eye symptoms and clinical signs were prevalent in the glaucoma patients with much worse tear film stability and more ocular surface staining than controls. Also there was an increase in severity of ocular surface illness with the number of drugs and daily instillations of eye drops, demonstrating the cumulative effect of chronic glaucoma management on ocular surface health.^35^

The observed results are also consistent with some previous study in which the authors reported that around 61% of glaucoma patients treated with conventional anti-glaucoma drugs had aberrant OSDI ratings with severe symptoms seen in roughly one quarter of patients. Higher OSDI scores were substantially linked with longer duration of therapy, numerous medicines, and preservative exposure and ocular surface abnormalities, suggesting that growing ocular surface damage is accompanied by increased patient-reported symptoms.^36^

In a systematic review and meta-analysis, Ghenciu et al.^32^, primarily focused on diabetes mellitus and not on glaucoma, but reported that patients with type 2 diabetes had significantly higher OSDI scores than non-diabetic controls and also had deterioration of other tear film parameters. This affirms the observation in the current study, that diabetes mellitus with coexisting glaucoma can cause additional aggravation of ocular surface symptoms.

The independent samples t-test revealed a statistically significant difference in the mean TBUT between the two groups (p = 0.003) in the present study. Also, the Chi-square analysis revealed a highly significant correlation between diabetes status and the clinical categories of TBUT (χ^2^ = 17.910, p < 0.001). Thus, the results show that DM has a significant effect on tear film stability in glaucoma patients living with DM. The mean TBUT of the glaucoma patients living with DM was 11.3 ± 9.0 seconds, which was lower than that of glaucoma patients living without DM (16.0 ± 10.6 seconds), showing that tear-film stability was reduced in the presence of diabetes. All the patients had glaucoma, and this data indicates that diabetes further deteriorates the ocular surface beyond the effects of glaucoma and chronic use of topical antiglaucoma drugs.

A considerable decline in ocular surface characteristics in individuals with glaucoma who are on topical medical therapy was also reported by a prospective investigation which showed reduced tear film stability, increased ocular surface staining, increasing dry eye symptoms and lower lipid layer thickness in glaucomatous patients compared to healthy controls.^37^

Other authors also provided a complete analysis of ocular surface illness related with topical glaucoma medication, further supporting the current findings. Their analysis indicated that individuals on long-term topical anti-glaucoma drugs often experience tear-film instability, decreased tear break-up time, ocular surface irritation and dry eye symptoms. They also noted that the severity of ocular surface illness increased with the number of drugs taken, length of treatment, and exposure to preservatives such as benzalkonium chloride.

Although their review did not specifically compare glaucoma subjects with and without diabetes mellitus or report independent t-test or Chi-square statistics, it synthesised evidence from multiple clinical studies showing statistically significant reductions in TBUT among glaucoma patients receiving chronic topical therapy. These findings are in line with the current study in that diabetes in addition to an ocular surface already damaged by glaucoma medication may further aggravate tear film instability.^38^

The independent samples t-test (*p = 0*.*003*) and Chi-square analysis (χ2 = 17.910, *p < 0*.*001*) strongly indicate that diabetes mellitus considerably affects tear film stability among glaucoma subjects in Owerri. These findings are in agreement with a meta-analysis, which has shown that diabetes and chronic glaucoma treatment independently influence tear film stability. The current study adds to the literature by revealing that diabetes causes an added deteriorating effect on TBUT in a glaucoma population, highlighting the need of regularly assessing tear film stability in the management of glaucoma patients with diabetes.

The decrease in TBUT found in glaucoma subjects living with diabetes could be physiologically justifiable. Chronic hyperglycemia causes oxidative stress, inflammation, ocular nerve dysfunction and loss of goblet-cells leading to diminished mucus production and instability of the tear film. In glaucoma patients, these alterations associated to diabetes are compounded by preservative toxicity and prolonged exposure to topical ocular hypotensive medicines, which increase tear film evaporation and decrease tear stability. Thus, the presence of these factors may be responsible for the much worse TBUT in glaucoma subjects living with diabetes than those without diabetes.

However, when the OSDI scores were categorized to normal, mild, moderate and severe categories, the Chi-square analysis yielded χ^2^ = 7.584 and p = 0.055. The p-value was slightly over 0.05 and the connection between diabetes status and categorical OSDI severity was therefore not statistically significant. Thus, the continuous OSDI scores were statistically substantially different between the two groups while the categorical distribution was not statistically significant in difference. This apparent disparity can be explained by the loss of information and statistical power that results when continuous scores are converted into broad severity categories. Subjects with differing OSDI scores may be categorized together, which might limit the power of the Chi-square test to identify modest but clinically important differences between groups.

These results are consistent with the findings of a previous study which investigated the factors associated with OSDI severity in a group of treated glaucoma patients, whereas the current study compared glaucoma subjects with and without diabetes mellitus. Both studies found statistically significant differences in ocular surface symptoms. The substantial associations found between various glaucoma and benzalkonium chloride exposure and OSDI severity suggest that several ocular and treatment-related variables impact OSDI ratings in glaucoma patients. In the current study, diabetes mellitus was another systemic component as shown by the significant independent sample t-test result of *p = 0*.*001*. However, the authors did not directly compare diabetic and non-diabetic glaucoma groups, nor did they reveal a Chi-square correlation between diabetes status and OSDI categories as in the present study.^36^

A prospective cross-sectional study in India which assessed glaucoma patients on chronic topical antiglaucoma drugs against controls using the OSDI questionnaire, Schirmer I test, tear break-up time, tear osmolarity and corneal fluorescein staining also support the results of the present study.The large OSDI difference (*p = 0*.*003*) observed by the authors aligns with the findings of the independent samples t-test of the present study (*p = 0*.*001*). (Ray et al.). Both studies report significant differences in mean OSDI scores. The agreement suggests considerable differences in ocular surface symptoms across subgroups of glaucoma patients according to their treatment exposure and related clinical features.

The theoretical standpoint of findings in the present are similar to the report of an evaluation on human clinical trials and experimental investigations on ocular surface illness caused or exacerbated by topical pressure reducing medicines. It was noted that toxic, inflammatory and allergic responses can be caused by active antiglaucoma drugs and preservatives, affecting the corneal and conjunctival epithelium, lacrimal and meibomian glands. These alterations can lead to tear film instability, superficial punctate keratitis, ocular pain, burning, redness and dryness, which increase OSDI scores and reduce treatment adherence and quality of life.^38^

Glaucoma patients living with diabetes mellitus had a higher OSDI score, which may be due to the combined effects of ocular surface dysfunction associated with diabetes mellitus and continuous use of antiglaucoma drugs. Diabetes mellitus may cause impaired corneal innervation, reduced corneal sensitivity, altered lacrimal gland function, changes in goblet-cell and meibomian-gland function, chronic inflammation, and epithelial abnormalities. The ocular surface disturbance may be worse than that reported by glaucoma patients living without diabetes mellitus when these diabetes-related alterations are combined with the toxic and inflammatory effects of topical glaucoma medicines, particularly preserved formulations.

## 5. Conclusion

Diabetes mellitus in glaucoma patients is associated with more deterioration of objective and subjective ocular surface function. The outcome of the present study shows that diabetes mellitus is an additional risk factor for ocular surface disease in glaucoma patients. The combination of glaucoma and diabetes may result in more rapid tear film instability, increased ocular pain, and decreased quality of life and adherence to long-term glaucoma medication. The findings underline the need of a thorough assessment of the ocular surface as an integral part of the normal management of glaucoma, particularly in patients with diabetes mellitus. The study consequently provides evidence for regular ocular surface examination in clinical eye care practice to allow for early identification, timely management and improved visual outcomes in those living with both glaucoma and diabetes mellitus.

## Data Availability Statement

The raw data supporting the findings of this study cannot be shared publicly due to institutional ethical restrictions regarding participant privacy.

## Funding

This research did not receive any specific grant from funding agencies in the public, commercial, or not-for-profit sectors.

## Declaration of competing interest

The authors declare that they have no known competing financial interests or personal relationships that could have appeared to influence the work reported in this paper.

## Acknowledgements

The authors acknowledge Prof. Emmanuel C. Esenwah for his support and guidance through the study.

## Notes

### Competing Interest Statement

The authors have declared no competing interest.

### Author Declarations

Ethical Approval was obtained from the Ethics Committee, School of Health Technology, Federal University of Technology, Owerri, to carry out this study after review, on the 8th of July, 2025. No specific protocol number was assigned by the committee. Informed consent was also gotten from subjects before inclusion in the study.

### Summary of Updates

The author names have been rearranged in this order: Chigozie Mary Chukwuoha,1 Emmanuel C. Esenwah,1 Nwakaego C. Ikoro,1 Godwin-Ovenseri-Ogbomo,2 Young Christian Azuamah,1 Nkiru Euphresia Odimegwu,3 Anthony U. Megwas,1 Genevieve Ugwoke,1 Jacqueline E. Obioma-Elemba,1 Yadirichukwu Eronini,4 Ezinne Chinenye Nkeremuzor1

